# Thigh muscle mass is associated with circulating valeric acid in healthy male volunteers

**DOI:** 10.1101/2023.08.11.23293978

**Authors:** Eva M. Hassler, Gernot Reishofer, Harald Köfeler, Wilfried Renner, Deutschmann H Hannes, Harald Mangge, Markus Herrmann, Stefan L. Leber, Felix Gunzer, Gunter Almer

## Abstract

Short-chain fatty acids (SCFAs) are circulating metabolites generated by gut microbiota. Recently, a link between age-related muscle loss and gut microbiota has been described, and SCFAs could play a mediating role in this relationship. Acetic, propionic, and butyric acid are the abundant forms and are considered to have health benefits, less is known about valeric and caproic acid.

In a cross-sectional study including 155 healthy volunteers, we investigated the relationship between muscle area, as well as intramuscular adipose tissue measured by magnet resonance imaging and circulating SCFAs. SCFAs and additional parameters were measured from serum and sport activities recorded by means of a questionnaire.

We found a significant positive association between valeric acid (VA) levels and the thigh cross-sectional muscle area in males. This relationship was independent of age, BMI and weekly training times and was not observed in females. No associations between other SCFAs and the thigh muscle cross-sectional area were observed.

To our knowledge, this is the first human study demonstrating a significant relationship between thigh muscle mass and VA, supporting the thesis of the “gut–muscle axis” with VA as a possible interacting player, on the one hand, affected by sex differences, on the other.

## Introduction

A balanced diet and regular exercise are the cornerstones of a healthy lifestyle and still the best protection against numerous diseases on a worldwide rise, such as cardiovascular (e.g. strokes and heart attacks) or metabolic diseases (e.g. type 2 diabetes) (1, 2). In recent years, clinicians have also recognized the importance of the gut microbiome as a metabolical key player, as well as its close relationship with obesity and related cardiometabolic disorders, specifically when the microbiome is out of balance (3, 4).

One of the main groups of metabolites produced by the gut microbiome are short-chain fatty acids (SCFAs). The main origin of all SCFAs is the gut, where they are produced in the large intestine by colon microbiota fermenting indigestible food components such as fibers like complex carbohydrates and resistant starch (5, 6). They are later metabolized in colonic epithelial cells, liver and other tissues, to serve as an energy source for the body, including the muscles (7).The three most common SCFAs are acetate, propionate, and butyrate (8). All three were found to have diverse physiological functions, for instance, affecting the blood lipid profiles, vitamin concentrations and energy production (9) as well as appetite, cardiometabolic health maintenance (10) and insulin sensitivity (11). Therefore, they are not only important for intestinal health (12), but because they enter the systemic circulation, they affect metabolism and peripheral tissue functions. Furthermore, they can alter dendritic cell and macrophage function to reduce the production of pro-inflammatory cytokines and they are also involved in the regulation of glycemic responses and triglyceride metabolism (6, 7).

SCFAs are very soluble and are absorbed easily from the intestinal lumen into the blood circulation via the portal vein. Acetic acid is usually the major short-chain fatty acid in human blood circulation and was found to be a possible energy source for the liver (13, 14). Butyric acid was found to be particularly of benefit for colon health as primal energy source for colonocytes (12). A very recent work by Lv et. al. showed an impact of serum levels from gut microbiota generated butyrate on the muscle mass index in healthy menopausal women (15). Furthermore, there are studies which demonstrate that SCFAs directly influence muscle mass *in vivo* (16-18). For example, Gao et al. demonstrated that chronic supplementation with butyrate preserves skeletal muscle in relation to the total body fat mass in rodents when incorporated into a high-fat diet (17). Also, Lahiri et al. clearly demonstrated the influence of gut microbiota on gene expression in muscle tissue and transcription of genes associated with skeletal muscle growth and function in mice (18).

All SCFAs show age related changes (19), but compared to “the big 3” – acetic (C2), propionic (C3) and butyric acid (C4) – valeric (C5) and caproic acid (C6) are commonly present at lower concentrations in feces and the circulation, and have not been considered as SCFAs strictly produced by the microbiota until now (19). However, in a recent article, they were found to be potent class I histone acetylase inhibitors (20). Valeric acid (VA) was further shown to inhibit the growth of *Clostridium difficile* when it was restored by fecal microbiota transplantation (21). In another preclinical study in mice, VA was found to reduce toxic injuries that were caused by radiation (22). However, compared to the other SCFAs, very little is known about the effects of VA on intestinal health and on health in general.

Some studies have shown that there is a link between the microbiome and age-related muscle loss, known as the “gut–muscle axis” (23-25). It is known that the pathophysiology of age-related muscle loss could be influenced by gut microbiota through different mechanisms of chronic inflammation and anabolic resistance (26). Loss of muscle mass e.g. with ageing or inactivity could lead to an increased fatty infiltration of the muscle tissue and therefore to an increased amount of intramuscular adipose tissue (IMAT) (27, 28). IMAT is also associated with a loss of muscle function (27). Furthermore, a negative effect of the amount of IMAT on glucose metabolism has been shown and several authors found relations between IMAT in the thigh muscles and different factors of cardiometabolic risk (29-32). Moreover, it was suggested by Goodpaster et. al. that too much IMAT may lead to an elevated free fatty acid (FFA) concentration in the skeletal muscles, which could cause a reduction in intramuscular blood flow followed by lower insulin diffusion capacity (33).

To achieve an accurate and radiation-free quantification of muscle mass and IMAT, magnet resonance imaging (MRI) is the most suitable method (29, 34, 35). Due to the development of new scanning techniques, examinations can nowadays be carried out in reasonable time (36, 37).

In the present study, we investigated a possible relationship between SCFAs, muscle mass and intramuscular adipose tissue in our study cohort of healthy active volunteers. Since other studies have shown associations between the gut microbiome and age-related muscle loss in men (24), we further wanted to investigate sex differences as well as relationships with body composition. Additionally, we compared the measured parameters between information on endurance and strength training.

## Materials and Methods

### Subjects

The included study subjects all gave written informed consent. The study participants were recruited from 12th of April 2018 until the 2nd February of 2019, and the study was approved by the local Ethics Committee of the Medical University Graz; (approval number: EK-Nr. 29-585 ex 16/17). The study had a cross-sectional design and 155 volunteers (78 females and 77 males) were included in the study. Some of the samples and data from our MRI-adipose tissue measurement study have been used and already published (38, 39). We excluded volunteers with known hyperlipidemia, metabolic disorders such as diabetes, and neurological and muscle diseases. Additionally, metallic implants and/or claustrophobia were reasons for study exclusion to MRI examinations. We collected parameters such as the height, weight, waist and hip circumference in every volunteer. We calculated ratios such as the waist/hip ratio and waist/height ratio.

The participants was given a questionnaire to note frequency, duration and nature of their sport activities as well as their lifestyle and dietary style (see supplements S1). The training frequency and duration were divided according to different training modes, in both endurance and resistance training. At the time of study inclusion, all subjects were healthy, and no one reported taking any antibiotics.

### Magnetic resonance imaging (MRI) and image processing

We performed MRI examination of the thigh after standardized positioning of a marker using a 3 Tesla magnetic resonance scanner (Siemens MAGNETOM® Prisma, Siemens Healthcare, Erlangen, Germany). The marker enables an exact localization of the measurements by using a standardized method developed by Mueller et al. (40). The MRI protocol consists of a standard 2-Point DIXON Sequence (TR 4.66ms, TE1 1.24ms, TE2 2.47ms, Flip angle 9°, base resolution 288, ISO Voxel Size: 1.4 x 1.4×1.4mm, PAT 4).

MRI data were analyzed using FIJI, which is an open-source image-processing package based on ImageJ (41). We used the image slice with the visible marker position for quantitative segmentation. The cross-sectional muscle area (MA) and intramuscular adipose tissue (IMAT) compartment were evaluated separately. The MA and percentage of IMAT in between the MA were calculated (38). Figure 1 shows segmentation results for and muscle area.

**Fig. 1.**
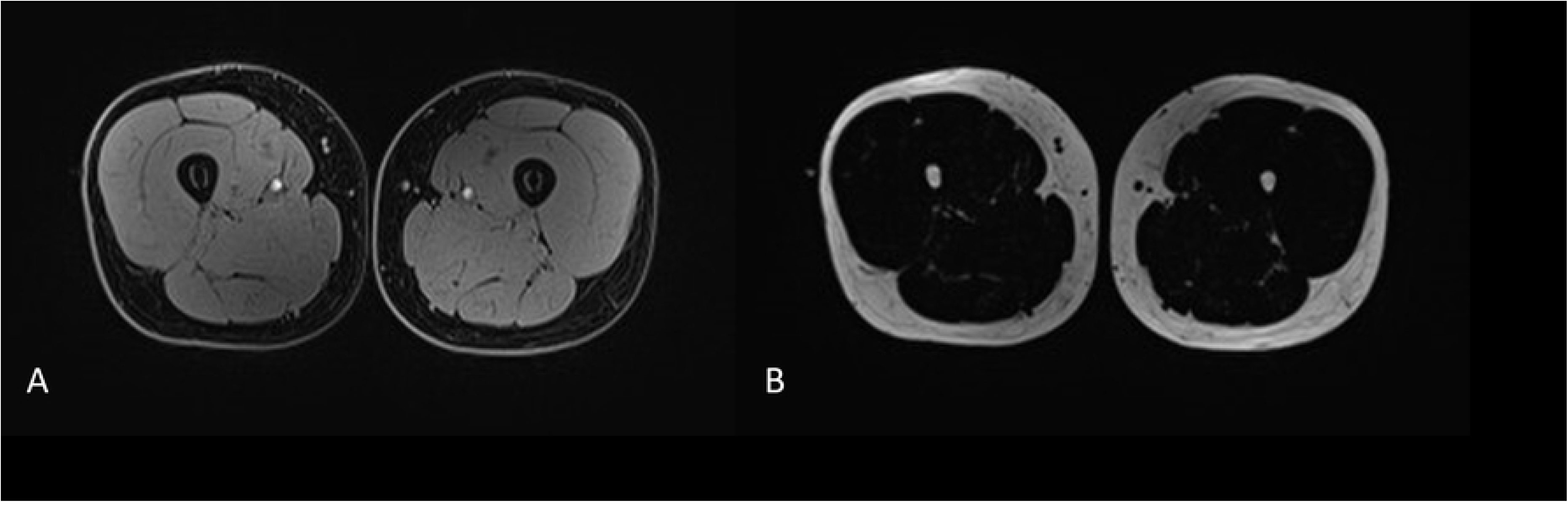
DIXON-MRI images of the mid-thigh. The cross-sectional muscle area was calculated from the water images (A) and the adipose tissue compartments were segmented from the fat images (B).

### Blood collection and clinical laboratory analysis

Venous blood samples were collected from every subject prior to MRI measurements. The blood samples were collected after a 12 h fasting period. Serum lipid panel contained total blood cholesterol (mg/dL), HDL (mg/dL), LDL (mg/dL), and serum triglycerides (mg/dL). The total cholesterol/HDL quotient was calculated. Serum levels of leptin (ng/ml) were analyzed using an appropriate human ELISA Kit® (BioVendor, Czech Republic) according to the manufacturer’s instructions. Standards, controls (high and low) and samples were measured in duplicates. The standard curves were calculated using the four-parameter algorithm. All blood samples were analyzed in the same laboratory with a standardized protocol using frozen serum (− 80 °C).

### Determination of short chain fatty acids by gas chromatograph-electron impact-mass spectrometer (GC-EI/MS)

Propionic acid-d6 was purchased from EQ Laboratories (Augsburg, Germany) and all other chemicals were supplied from Sigma Aldrich (St. Louis, MO, USA). A measure of 10 µl of an internal standard mix was added to 100 µl of serum samples, resulting in a final concentration of 100 µM acetic acid-d4, propionic acid-d6, butyric acid-d8, valeric acid-d9 and hexanoic acid-d3 each. Subsequently, 110 µl of methyl tert-butyl ether (MTBE) was added. The mixture was shaken for 10 min and then centrifuged in a lab centrifuge at 2500 rpm. The upper phase was transferred into a GC–MS vial. The injection volume was 1 µl in splitless mode at 250°C injector temperature. The GC oven was equipped with a 1 DB-WAX UI GC-column (30m x 0.25mm ID x 0.25µm film) and an initial oven temperature of 40°C was held for 2 min. Thereafter, it was ramped up to 150°C at a rate of 15°C/ min followed by an increase of 5°C/min up to 170°C and an increase of 20°C/min up to 250°C, where the temperature was finally held for 5 min. Helium was used as a carrier gas at a flow rate of 1.3 ml/min. The transfer line temperature was set to 280°C. The mass spectrometer was operated in SIM mode using the following m/z values: 60, 63, 73, 74, 76, 79 and 80 at their respective retention times. Acetic (C2), propionic (C3), butyric (C4), valeric (C5) and hexanoic (C6) acid were determined by one-point calibration versus their corresponding internal standards.

### Statistics

Statistics were performed using SPSS version 27 (IBM). Group differences between males and females in the descriptive evaluation have been performed using Welch’s *t*-test. Statistical comparison of parametric continuous subject characteristics was accomplished using the two-sided Student’s *t*-test with Pearson’s linear correlation coefficients over the whole study population and separated into groups according to sex. The distribution was tested for normality using the Shapiro–Wilk test. A p-value <0.05 was considered to be statistically significant. A stepwise multiple linear regression analysis was performed to adjust the relations for age, BMI, training activities and serum lipid panel including Fisher’s z-transformation for the Pearson correlation coefficients.

## Results

### Subjects

The study had a cross-sectional design including 155 volunteers; 78 females/77 males; age: min 21.4-max 76.3 (males 41.65 ± 11.53/mean ± SD, females 45.31 ± 10.41/mean ± SD). Table 1 shows the age, BMI, waist/hip and waist/height ratios, muscle areas and areas of adipose tissue compartments including the ratios of the fat compartments, respectively, and serum levels of SCFAs, as well as the lipid profile of the entire study population. The parameters were indicated for both males and females.

**Table 1:** Descriptive statistics of the study population.

| Anthropometric measurements | Total (n=155) |  | Female (n=78) |  | Male (n=77) |  | p-value |
| --- | --- | --- | --- | --- | --- | --- | --- |
| age (years) | 43.47 | ± 11.10 | 45.31 | ± 10.41 | 41.65 | ± 11.53 | 0.040* |
| BMI kg/m<sup>2</sup>) | 23.74 | ± 3.12 | 22.88 | ± 3.29 | 24.60 | ± 2.70 | **0.001\*\*\*** |
| Waist/hip ratio | 0.82 | ± 0.07 | 0.78 | ± 0.06 | 0.86 | ± 0.07 | **<0.001\*\*\*** |
| Waist/height ratio | 0.47 | ± 0.05 | 0.46 | ± 0.06 | 0.48 | ± 0.05 | **0.009\*\*\*** |
| **Compartments and ratios** |  |  |  |  |  |  |  |
| Muscle cross-sectional area (px) | 14253.08 | ± 3395.47 | 11702.30 | ± 1486.93 | 16771.17 | ± 2818.97 | **<0.001\*\*\*** |
| SAT/(SAT+IMAT) | 0.88 | ± 0.05 | 0.89± | ± 0.05 | 0.87± | ± 0.05 | **0.007\*\*** |
| IMAT/MA | 0.06 | ± 0.05 | 0.08 | ± 0.05 | 0.04± | ± 0.02 | **<0.001\*\*\*** |
| **Laboratory measurements** |  |  |  |  |  |  |  |
| Leptin | 8.46 | ± 8.73 | 9.15 | ± 9.20 | 7.68 | ± 8.18 | 0.312 |
| **Lipid profile** |  |  |  |  |  |  |  |
| Total blood cholesterol (mg/dl) | 194.81 | ± 32.49 | 200.18 | ± 30.15 | 189.51 | ± 34.02 | **0.040\*** |
| HDL-C (mg/dl) | 68.97 | ± 17.78 | 76.10 | ± 18.74 | 61.94 | ± 13.60 | **<0.001\*\*\*** |
| LDL-C (mg/dl) | 102.93 | ± 30.84 | 102.74 | ± 29.37 | 103.10 | ± 32.36 | 0.943 |
| Triglycerides (mg/dl) | 115.56 | ± 88.95 | 108.17 | ± 99.68 | 122.86 | ± 76.87 | 0.306 |
| **SCFA** |  |  |  |  |  |  |  |
| C2 (μmol/L) | 10700.92 | ± 9583.07 | 10207.33 | ± 8810.55 | 11188.19 | ± 10239.52 | 0.523 |
| C3 (μmol/L) | 572.33 | ± 353.33 | 566.44 | ± 335.27 | 578.13 | ± 372.39 | 0.838 |
| C4 (μmol/L) | 56.51 | ± 32.43 | 54.50 | ± 30.00 | 58.49 | ± 34.76 | 0.446 |
| C5 (μmol/L) | 11.77 | ± 7.83 | 11.07 | ± 7.36 | 12.48 | ± 8.06 | 0.257 |
| C6 (μmol/L) | 60.52 | ± 40.72 | 58.14 | ± 38.96 | 62.86 | ± 42.51 | 0.472 |
| **Sport profile** |  |  |  |  |  |  |  |
| Endurance (min/w) | 221.20 | ± 371.15 | 188.65 | ± 403.77 | 253.33 | ± 335.50 | 0.280 |
| Resistance (min/w) | 87.00 | ± 105.18 | 67.18 | ± 85.88 | 106.56 | ± 118.59 | **0.019\*** |
| Endurance (Sessions/w) | 2.72 | ± 2.12 | 2.59 | ± 2.22 | 2.84 | ± 2.01 | 0.455 |
| Resistance (Sessions/w) | 1.71 | ± 1.86 | 1.41 | ± 1.70 | 2.01 | ± 1.97 | **0.044\*** |

*Sex differences were evaluated via t-test. The shown p-values mark significant sex differences (values are reported as mean ±SD). Intermuscular adipose tissue (IMAT); subcutaneous adipose tissue (SAT); low-density lipoprotein (LDL); high-density lipoprotein (HDL); p < .05 indicated as \*, p < .01 indicated as \*\*, p < .001 indicated as \*\*\**

The BMI was statistically significantly higher in the male group than in females. The muscle cross-sectional area showed significantly higher values in the male group than in females. The percentage of IMAT per MA was significantly lower in the male group. HDL was higher in females. No statistically significant differences were found between the weekly endurance training times in men and women, but men performed significantly longer weekly resistance training units. Comparing the levels of all SCFAs between both sexes, no significant differences were found between men and women (Table 1).

### Pearson correlations differ in males vs. females

Table 2 shows the correlation of SCFAs, adipose tissue and muscle tissue segmentation results. The most interesting observation in this study was a significant correlation of the cross-sectional muscle area of the mid-thigh with circulating VA (C5) in healthy males (r=0.324/p-value=.004). This association was not found in females (r=-.030/p-value=.793). The scatterplots of VA versus muscle mass are shown in Fig.2. No correlation was found between circulating SCFAs and the weekly duration of resistance training in both males and females.

**Fig. 2.a.**
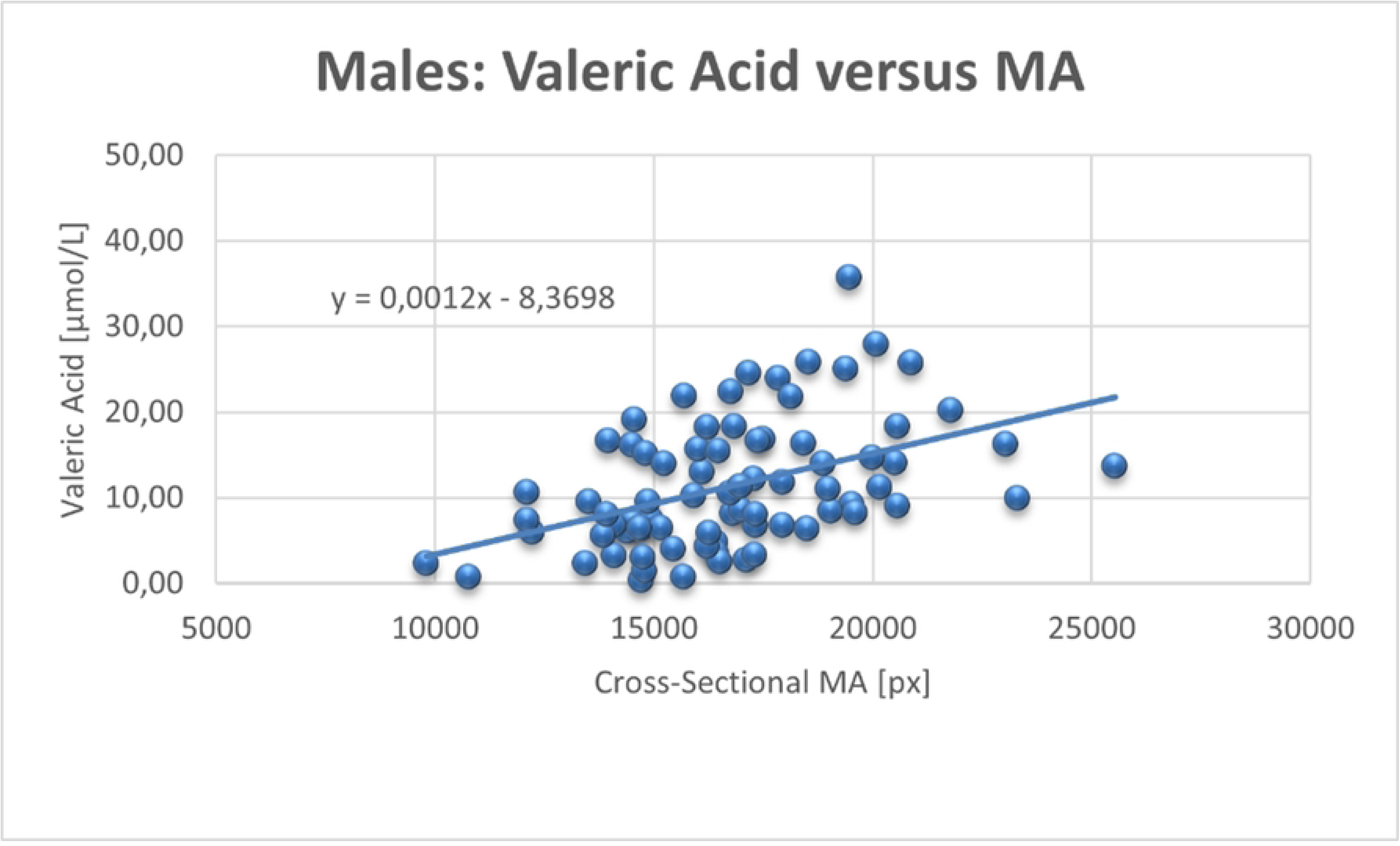
Scatterplot of the correlations between valeric acid (VA) and muscle area (MA) including regression line in males

**Fig. 2.b.**
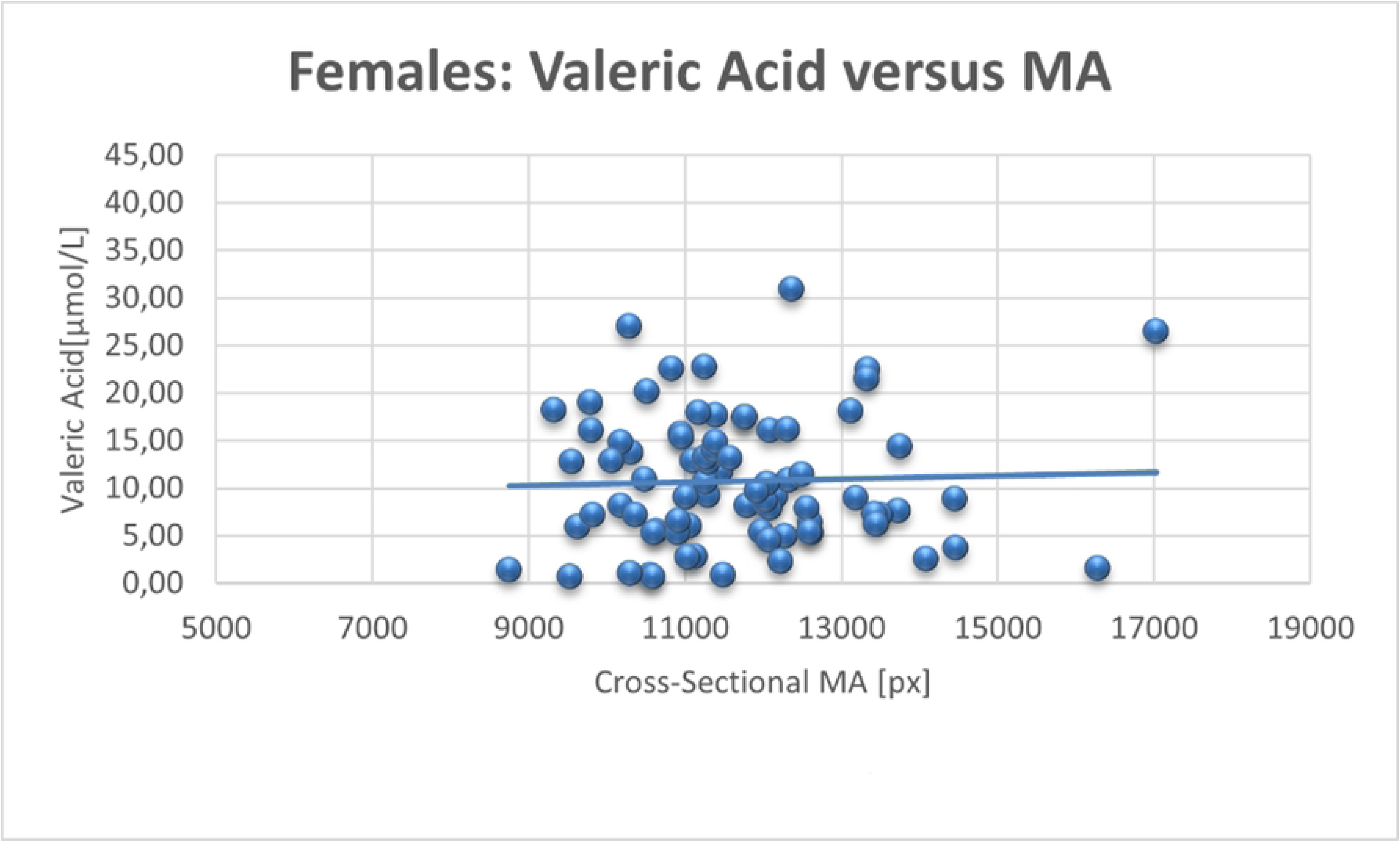
Scatterplot of the correlations between valeric acid (VA) and muscle area (MA) including regression line in females.

**Table 2:**
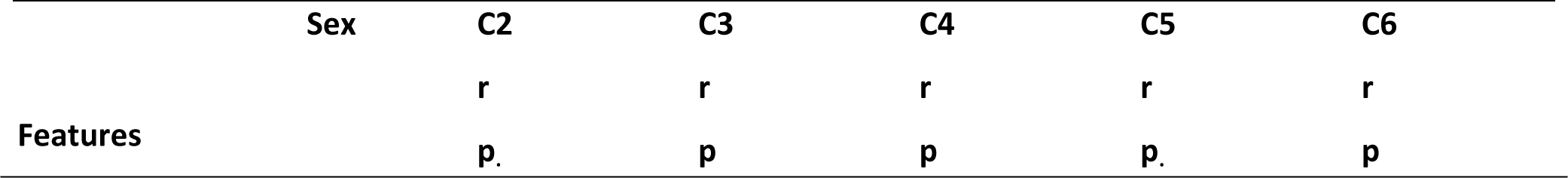
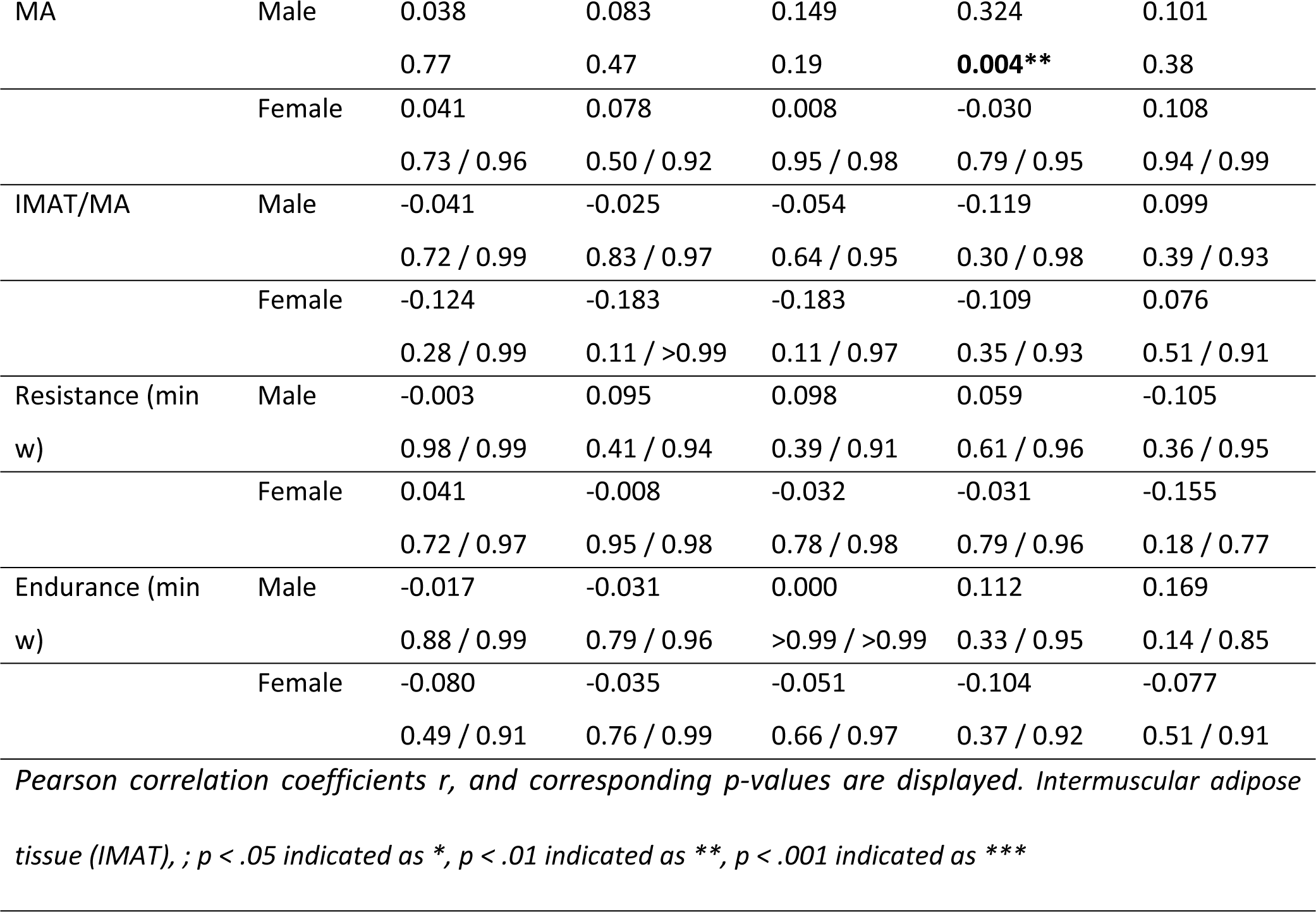
Pearson correlation results between SCFA serum levels, thigh MA, intermuscular ad tissue segmentation results, and weekly training times.

### Results of the multivariate regression analysis

In an additional multivariate regression analysis with VA (C5) as constant, we could show that the relationship between the cross-sectional muscle area and VA in males remains significant. The model was adjusted for age, BMI, endurance and resistance training times and serum lipids. In females, none of the factors are related or influenced the VA levels (Table 3. a, b).

**Table 3a:**
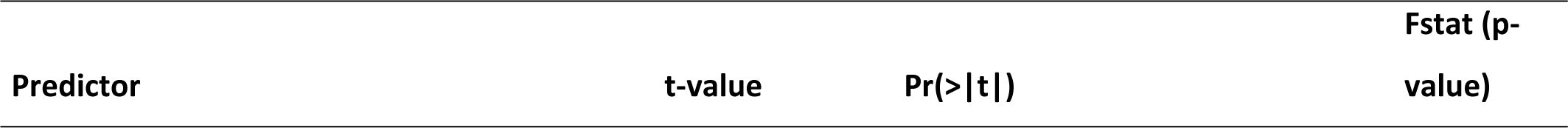
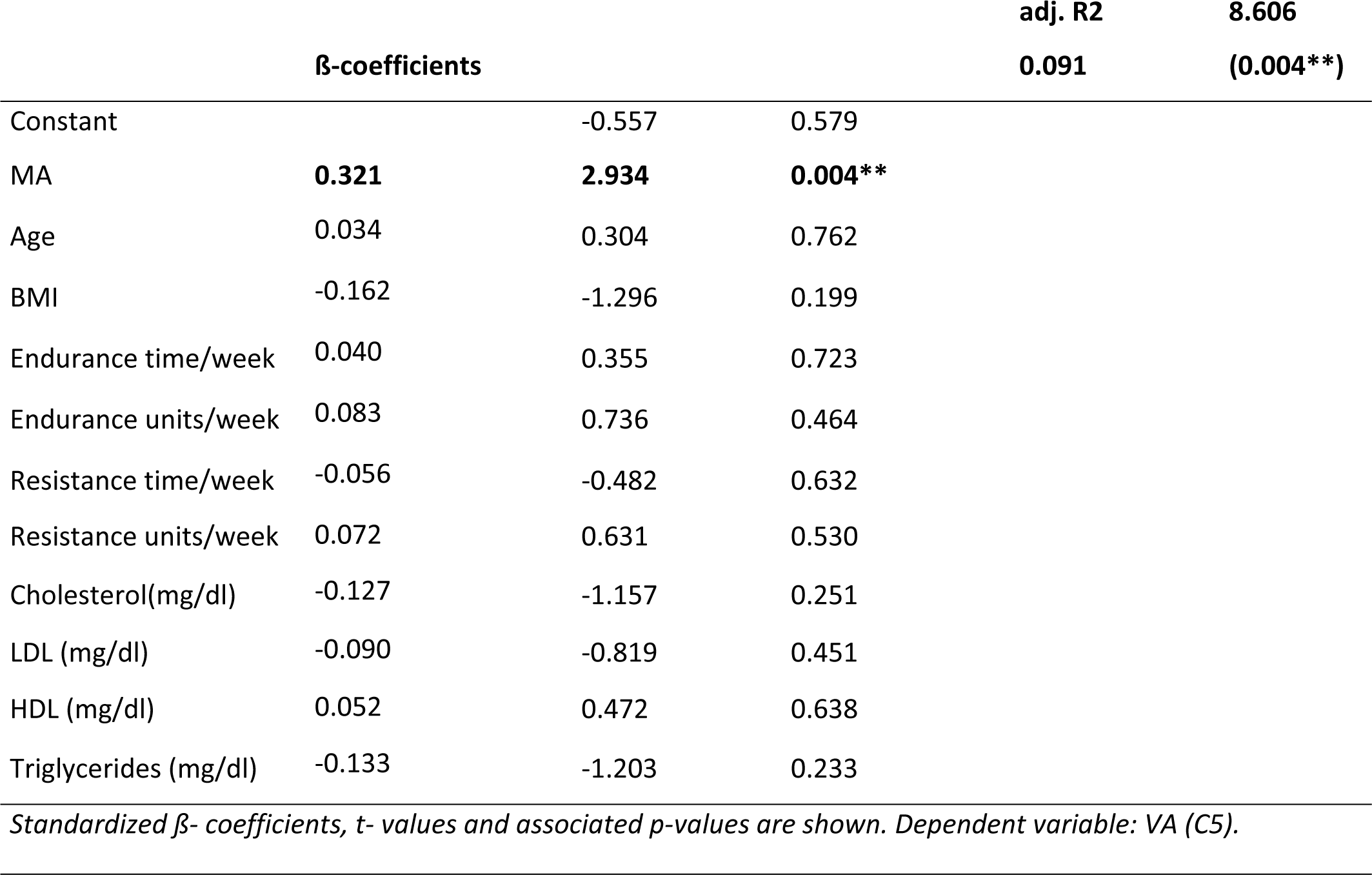
Effect of cross-sectional MA, age, body mass index, resistance and endurance training and serum lipid panel on VA levels in men (stepwise inclusion).

**Table 3b:**
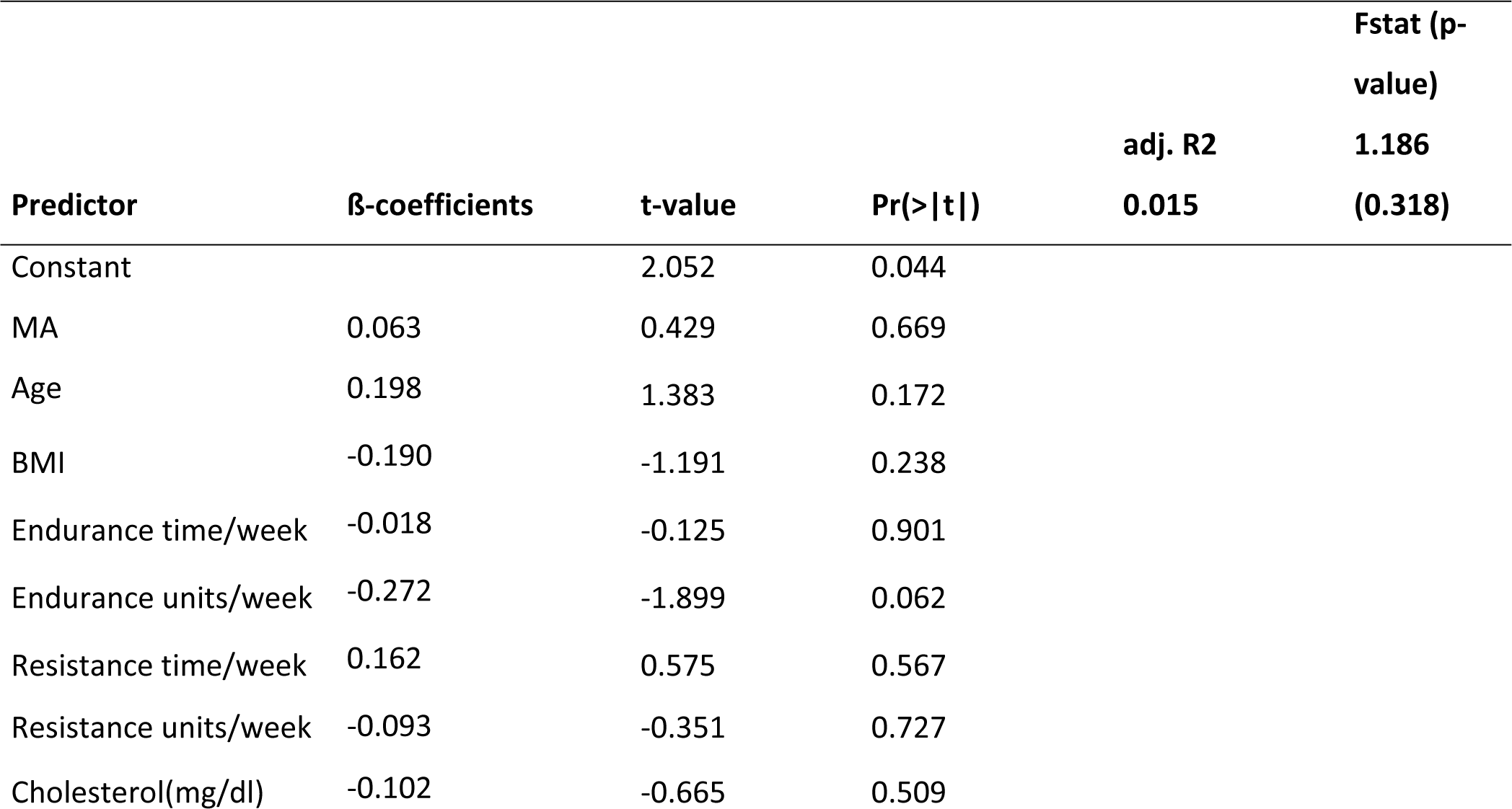
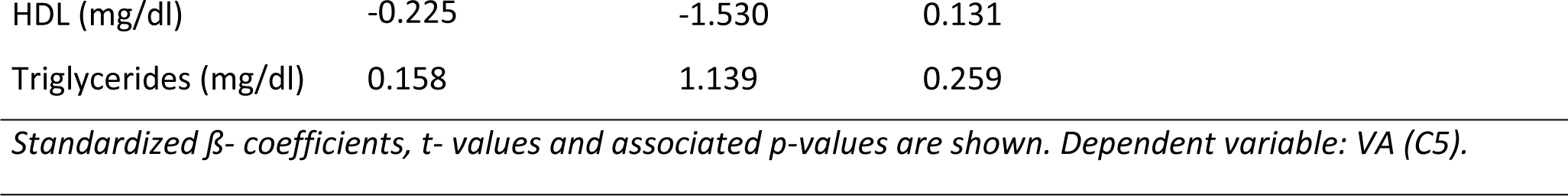
Effect of cross-sectional MA, age, body mass index, resistance and endurance training and serum lipid panel on VA levels in women (stepwise inclusion).

|  |  |  |  | adj. R2 | Fstat (p-value) |
| --- | --- | --- | --- | --- | --- |
|  |  |  |  | 0.015 | 1.186 (0.318) |
| Predictor | β-coefficients | t-value | Pr(>\|t\|) | | |
| Constant |  | 2.052 | 0.044 |  |  |
| MA | 0.063 | 0.429 | 0.669 |  |  |
| Age | 0.198 | 1.383 | 0.172 |  |  |
| BMI | -0.190 | -1.191 | 0.238 |  |  |
| Endurance time/week | -0.018 | -0.125 | 0.901 |  |  |
| Endurance units/week | -0.272 | -1.899 | 0.062 |  |  |
| Resistance time/week | 0.162 | 0.575 | 0.567 |  |  |
| Resistance units/week | -0.093 | -0.351 | 0.727 |  |  |
| Cholesterol(mg/dl) | -0.102 | -0.665 | 0.509 |  |  |

|  |  |  |  |
| --- | --- | --- | --- |
| HDL (mg/dl) | -0.225 | -1.530 | 0.131 |
| Triglycerides (mg/dl) | 0.158 | 1.139 | 0.259 |

*Standardized $\beta$ - coefficients, t- values and associated p-values are shown. Dependent variable: VA (C5).*

When the cross-sectional muscle area was used as the dependent variable in the model, again, VA was found to be a significant influencing factor along with BMI, age, weekly weight training duration, the number of weekly endurance training sessions, and HDL. Again, in the female group, there was no correlation with VA (Table 4. a, b).

**Table 4a:**
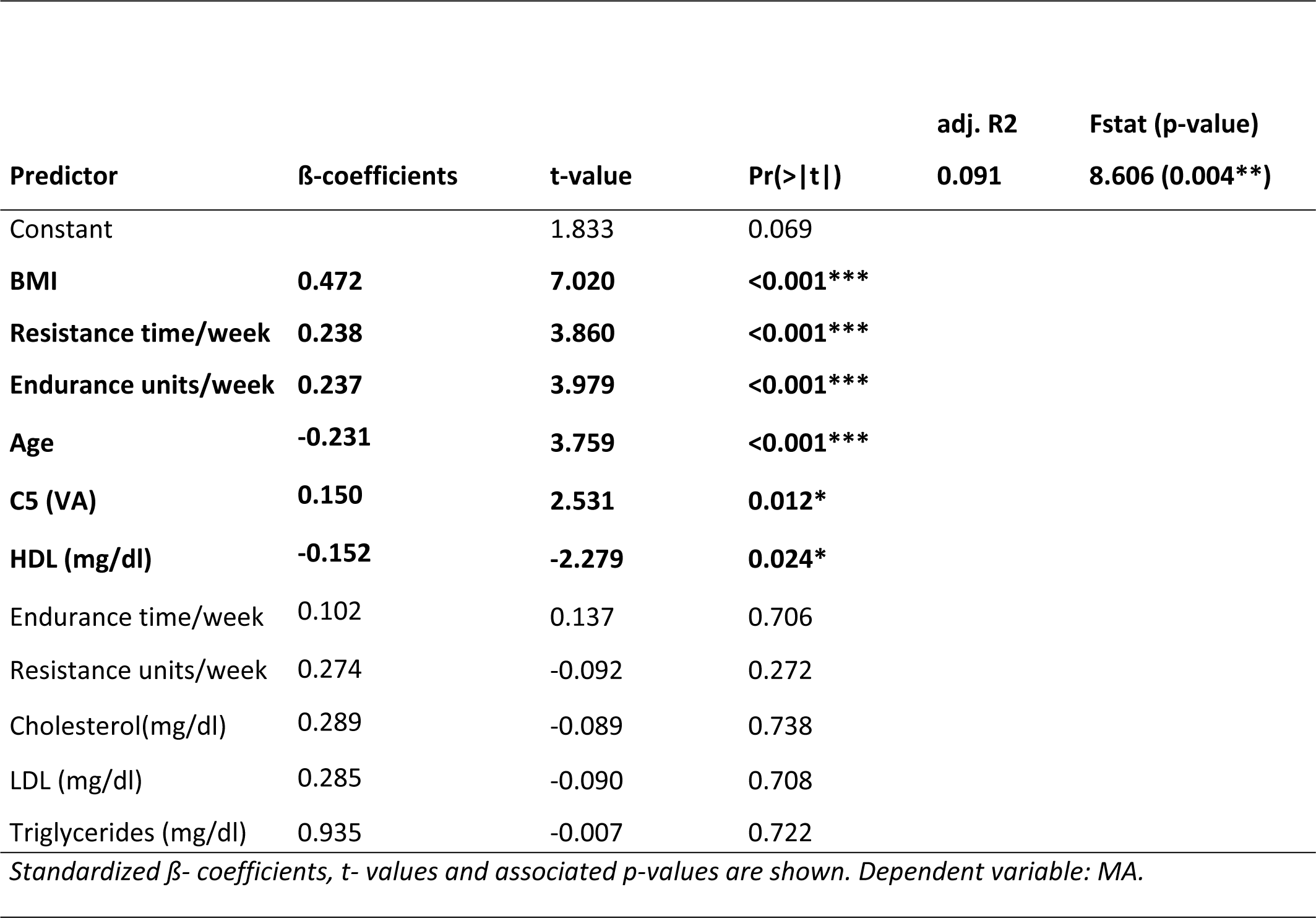
Effect of VA, age, body mass index, resistance and endurance training and serum lipid panel on cross-sectional muscle area in men (stepwise inclusion).

**Table 4b:**
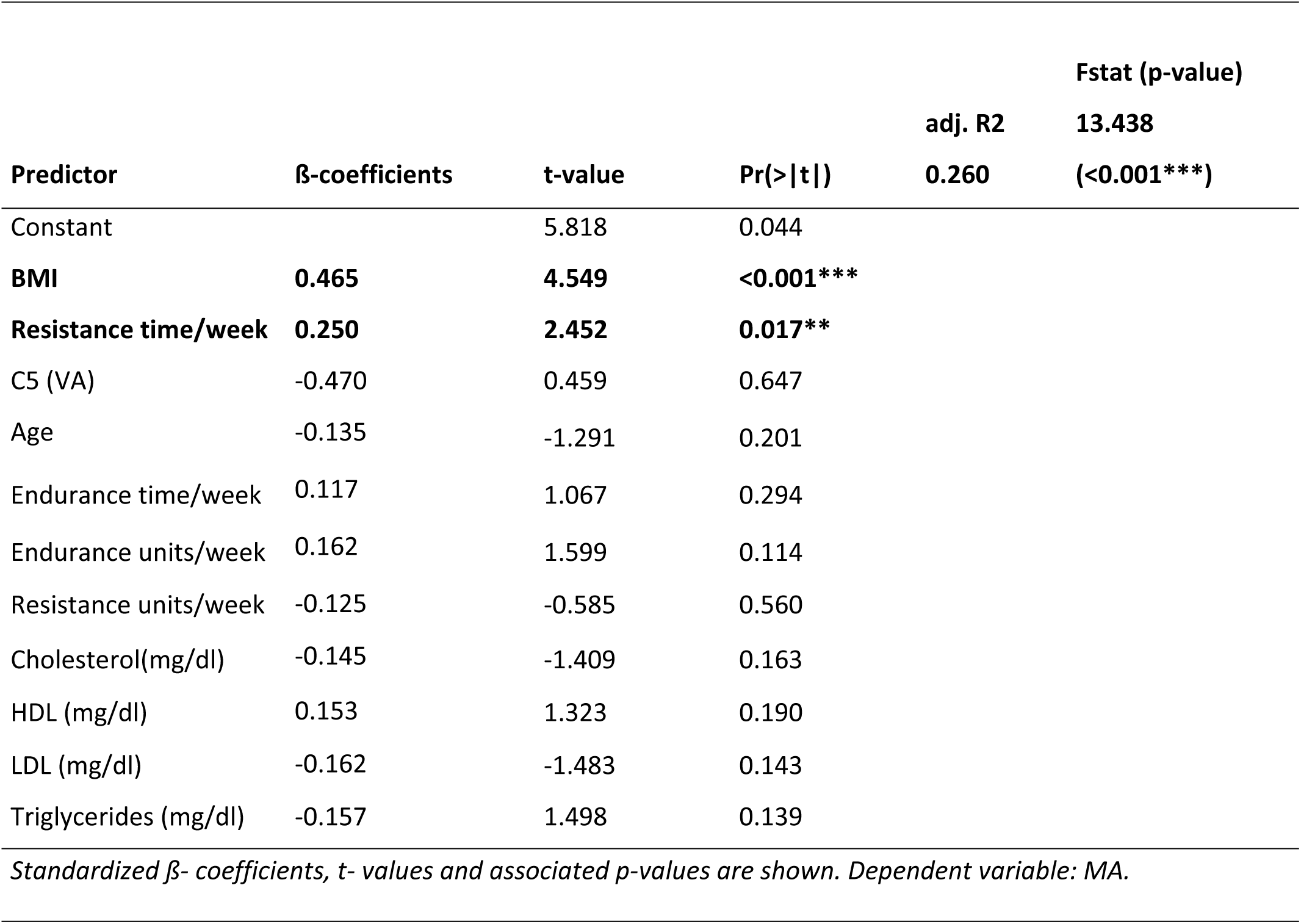
Effect of VA, age, body mass index, resistance and endurance training and serum lipid panel on cross-sectional muscle area in women (stepwise inclusion).

| Predictor | β-coefficients | t-value | Pr(>\|t\|) | adj. R2 | Fstat (p-value) |
| --- | --- | --- | --- | --- | --- |
|  |  |  |  | 0.260 | 13.438<br>(<0.001***) |
| Constant |  | 5.818 | 0.044 |  |  |
| **BMI** | **0.465** | **4.549** | **<0.001\*\*\*** |  |  |
| **Resistance time/week** | **0.250** | **2.452** | **0.017\*\*** |  |  |
| C5 (VA) | -0.470 | 0.459 | 0.647 |  |  |
| Age | -0.135 | -1.291 | 0.201 |  |  |
| Endurance time/week | 0.117 | 1.067 | 0.294 |  |  |
| Endurance units/week | 0.162 | 1.599 | 0.114 |  |  |
| Resistance units/week | -0.125 | -0.585 | 0.560 |  |  |
| Cholesterol(mg/dl) | -0.145 | -1.409 | 0.163 |  |  |
| HDL (mg/dl) | 0.153 | 1.323 | 0.190 |  |  |
| LDL (mg/dl) | -0.162 | -1.483 | 0.143 |  |  |
| Triglycerides (mg/dl) | -0.157 | 1.498 | 0.139 |  |  |

*Standardized $\beta$ - coefficients, t- values and associated p-values are shown. Dependent variable: MA.*

## Discussion

Evidence showing the metabolic effects of the physiologically lower concentrated SCFAs VA and CA is still lacking. To our knowledge, we are the first to show a significant association of the cross-sectional muscle area of the thigh with circulating VA. The second interesting observation is that it was only found in healthy males. Furthermore, the relationship remained statistically significant in a multivariate regression analysis with VA as the dependent variable adjusted for age and BMI. Neither the endurance or resistance training time and units per week showed a statistically significant relationship with VA (C5) in the multivariate regression model.

The cause-and-effect principle of our observation cannot be clearly answered at this stage. Different underlying pathophysiological mechanisms could explain the influence of the gut microbiome on skeletal muscle metabolism (18, 23). Therefore, we assume, that VA could influence muscle mass.

On the other hand, muscle mass may also have an influence on serum levels of VA. Several studies showed that exercise affects muscle mass, and the microbial diversity in the gut was suggested to be increased (42-44), involving mechanisms such as myokine release, increased intestinal transit, or the secretion of neurotransmitters and hormones, including the promotion of an anti-inflammatory state (43-46). Some animal studies using mouse and rat models indicated changes in the gut microbiome with an increase in several short-chain fatty acid (SCFA)-producing bacteria in mice after strength training (47, 48). A recent study on elite athletes indicated that the gut microbiota could influence their training performance. They found that *Veillonella atypica* was positively associated with increased physical performance by the metabolization of lactate into propionate in these high-performance athletes (49). A longitudinal study in humans describes changes in the gut microbiome after completing an 8-week endurance program (50). Here, no changes were found after an 8-week intervention with strength training.

Compared to these studies, we did not observe a significant influence of endurance or resistance training times on VA levels in males in a multilinear regression analysis, but although we did find a significant influence of training interventions on the cross-sectional muscle area. A reason for this could be the recording of our training modalities with a questionnaire, which may be seen as a study limitation in this context.

Recent research with mice found an association of gut microbiota dysbiosis with an underrepresentation of species involved in SCFA synthesis and reduced expression of free fatty acid receptors in enteroendocrine cells, leading to a negative effect for the glucose metabolism of skeletal muscle cells (51). Furthermore, a link between insulin sensitivity and circulating SCFAs has been described in human studies (11). In this context, studies demonstrated that the G protein-coupled receptor 41 plays a role in insulin responsiveness and is enhanced by both propionic and valeric acids on glucose uptake shown in 3T3-L1 adipocytes and C2C12 myotubes; furthermore, a valerate-induced increase in basal glucose uptake in C2C12 myotubes was found, suggesting a role for VA in muscle metabolism (52, 53).

Since the amount of intramuscular adipose tissue is also independently associated with insulin resistance, we would have expected a relationship between IMAT/MA and acetic or valeric acid in this context (11, 30, 31), but that was surprisingly not the case. Using the Welch’s test, we did not find significant differences in the VA levels between males and females, contrary to other studies (54). Nevertheless, we found an expected difference in the cross-sectional muscle area of the thigh between males and females.

However, our observation supports the findings of previous studies that have shown differences in the gut microbiome between sarcopenic and non-sarcopenic male subjects (23, 24). Compared to that study, we included healthy male and active subjects. The prior study, which found a causal effect between gut microbial synthesis of SCFA butyric acid and the appendicular muscle mass in postmenopausal healthy women (15), could not be supported by our results as we did not observe this in our female cohort. Nevertheless, the associations we observed between VA and MA in males suggest that muscle mass influences VA rather than *vice versa*, as we did not see this observation in females, possibly due to the lower average muscle mass.

By now, only some single studies used VA for therapeutical application. One animal study showed VA glyceride ester feeding to chicken broilers. It resulted in a positive influence of aspects of their performance and a reduction in the number of necrotic enteritis incidences (55). Interestingly, valproic acid, a branched SCFA derivate of VA, is already used as a therapeutic agent for spinal muscular atrophy (SMA). Its administration led to an increase in the expression of survival motoneuron (SMN) mRNA (56, 57). Furthermore, treatment with valproic acid resulted in an improvement in gross motor functions for SMA patients (57). Whether the intestinally produced valproic acid can interact with SMN protein levels and thereby have effects on, e.g., muscle mass, or what underlying mechanisms could be responsible for the relationship between muscle mass and VA observed in our male cohort are not currently clear and would need further investigation to ascertain.

There are some other limitations to this study. The sex hormone status was not recorded, which could play an important role in the build-up of muscle mass. Additionally, information about dietary habits or the use of antibiotics and probiotics was not optimally surveyed. Thus, the observed interrelationships should be investigated in further longitudinal studies. Due to the original study design, the gut microbiota compositions were not examined in this cross-sectional study. Further research is planned to examine sex-specific relationships between gut microbiota, enteral and circulating SCFAs, and changes after training interventions. In terms of disadvantages with the recording of sporting activities with a questionnaire, we intend to carry out more precise examinations. Therefore, studies with performance diagnostics could also be carried out in the future.

In conclusion, we were able to indicate a significant association between VA and thigh muscle mass in healthy males, which, to our knowledge, has not been described before. In relation to our investigation and other studies, we suppose that training interventions, which effect muscle mass could influence the gut microbiome and consecutively VA concentrations. On the other hand, also an influence of VA on muscle mass is possible, as prior studies found an effect of the gut microbiome on muscle metabolism. Nevertheless, our observation establishes a new view on VA, as it possibly has a more important role in energy metabolism than known and could be examined more closely in future studies.

## Data Availability

All relevant data are within the manuscript and its Supporting Information files.

## Abbreviations

CA: caproic acid (C6)
FFA: free fatty acids
GC-EI/MS: gas chromatograph-electron impact-mass spectrometer
IMAT: intramuscular adipose tissue
MA: muscle area
MRI: magnet resonance imaging
MTBE: methyl tert-butyl ether
SCFA: short-chain fatty acids
SMA: spinal muscular atrophy
SMN: survival motoneuron
VA: valeric acid (C5)

## Additional information

Competing financial and non-financial interests are disclosed by all contributing authors. This research received no external funding.

